# A wearable EEG device: LANMAO Sleep Recorder compared to polysomnography in terms of EEG recording and sleep analysis

**DOI:** 10.1101/2023.05.30.23290376

**Authors:** Xin Zheng, Guannan Xi, Huijie Lei, Dongbin Lyu, Yu Zhang, Chengmei Yuan, Anchen Gao, Siyang Huang, Jian Jiang

**Author notes:** Corresponding author: Jian Jiang, Shanghai Quanlan Technology Co., Ltd, No. 3666 Sichen Road, Sheshan Town, Songjiang District, Shanghai 201600, China.

## Abstract

**Background:** Polysomnography (PSG) is the gold standard for sleep monitoring and diagnosis, yet it is difficult to use in home environments. This study evaluated the performance of a wearable electroencephalographic (EEG) device, LANMAO Sleep Recorder in EEG recording and sleep staging algorithm by comparing with PSG.

**Method:** Sleep of 7 Chinese adults were recorded concurrently with PSG and LANMAO devices. First, we validated the consistency of the raw signals with relative spectral power and Pearson correlation coefficient. Second, we evaluated the performance of the automated sleep staging algorithm integrated in the LANMAO device by comparing with the staging by experts.

**Results:** The Pearson correlation coefficient between the relative spectral power of multiple frequency bands during the sleep stages ranged from 0.7613 to 0.8816, with the strongest correlation observed for delta waves (r=0.8816). The overall F1-Score of the automated sleep staging algorithm was 84.03%, with individual F1-Scores for each class as follows: Wake: 93.67%, REM: 87.23%, Light Sleep: 72.10%, and Deep Sleep: 82.82%.

**Conclusion:** The results suggest that the EEG recorded by the LANMAO Sleep Recorder is precise and valid, and its automated sleep staging algorithm can accurately perform sleep staging with high accuracy. Therefore, in specific scenarios such as the home environment, LANMAO devices can work as a promising PSG alternative for sleep monitoring.

## INTRODUCTION

Sleep disorders, such as insomnia, sleep apnea, and narcolepsy, represent a group of widely prevalent public health issues that are highly associated with mental disorders[1][2], and have widely adverse impact on individual performance, and society burden[3][4]. Polysomnography (PSG), a gold standard, is generally used to record and diagnose sleep disorders [5]. PSG can provide comprehensive monitoring of the patient’s sleep, by placing multiple electrodes on the patient, it provides information mainly from electromyography (EMG), electroencephalography (EEG), electrocardiography (ECG), and respiratory rhythm. However, PSG can increase the patient’s physical discomfort and psychological burden during sleep, which is not in line with the principle of sleeping comfortably emphasized in cognitive behavioral therapy for insomnia (CBT-I)[6]. Additionally, PSG is laboratory based, which is not suitable for long-term monitoring in home environments which is needed for the general population. What is more, it takes a lot of effort and time to complete sleep staging diagrams based on PSG data. The raw data from PSG requires a series of processing steps and analysis by sleep experts manually based on the American Academy of Sleep Medicine’s (AASM) Sleep Staging Manual[5]

To tackle some limitations of PSG, several lightweight and wearable EEG recording devices which can record users’ sleep EEG and perform sleep analysis in home environments have shown up[7][8][9]. For instance, prior studies indicated that single-channel EEG on the forehead can produce results comparable to PSG when evaluating parameters such as REM, N2, and N3 sleep stages [9][10]. Arnal and his colleagues[8] applied a newly developed headband-form EEG device, then using a neural network model (Long Short-Term Memory, LSTM[11]) to predict sleep stages, and the results showed a high accuracy comparing to PSG. In addition, previous studies indicated that the accuracy of automatic sleep staging algorithms based on machine learning or deep learning using PSG data can reach over 80% accuracy [12][13], which is close to the level of sleep experts. However, before the large-scale usage of home-based portable EEG devices by the general population, the accuracy and reliability of portable EEG devices and their automatic sleep scoring still need to be verified.

In the current study, we set out to validate a newly wearable sleep monitoring device, LANMAO Sleep Recorder (abbreviated as LM hereafter) in the Chinese general population. LM records the frontal single-channel EEG data during sleep and the sleep staging algorithm is based on the LightGBM framework[14][15][16], which exhibits high accuracy and computational speed. We aimed to 1) Compare real-time frontal EEG data collected by LM with the PSG FP1-FP2 channel EEG data, 2) Compare the sleep staging results produced by LM (the frontal EEG and head movement information) with PSG multi-channel data that analyzed by multiple sleep experts.

## METHODS

### Subjects

In total, 10 volunteers were recruited from the local community in Shanghai, China, through advertisement, including 7 males and 3 females whose ages range from 18 to 35. There are some exclusion criteria, including: 1) currently pregnant or lactating; 2) history of severe cardiac, neurological, or psychiatric disorders within the past 12 months; 3) morbid obesity (BMI ≥ 40); 4) recent history of drug dependence; 5) patients with narcolepsy or rapid eye movement sleep behavior disorder, to avoid data recording abnormalities; 6) skin problems such as acne, lesions, or allergic rash on the forehead.

Each participant provided at least one night of data, with a total of 10 nights data, excluding pre-experiment verification of equipment and procedures. Three participants were excluded due to the problems of electrode connection or poor signal quality, the final dataset consisted of one overnight recording from each of seven participants. It is worth noting that we did not exclude cases of mild or moderate sleep disorders during recruitment of participants. As the population with sleep disorders is the target population for insomnia treatment strategies such as CBT-I, collecting and comparing data on this population is of practical significance, so we did not exclude data with short total sleep time or fragmented sleep patterns. The final dataset consisted of one overnight recording from each of seven participants.

### Protocol

Potential participants first had a brief video call with the research staff, followed by a face-to-face interview, during which they needed to provide informed consent and complete a detailed questionnaire including personal information, health status, sleep status, lifestyle.

Once qualified and agreed to participate in the experiment, participants were accompanied by sleep experts to the sleep monitoring room at Shanghai Mental Health Center either Shanghai Center for Brain Science and Brain-Inspired Technology at their convenience, during which both PSG and LM devices were worn and recorded. The sleep monitoring rooms are designed to simulate a home sleep environment with beds, tables, chairs, and toilets. Participants need to arrive about half an hour before their scheduled sleep time, the start and end of the data recording were set based on the participants’ self-selected lights out and lights on time, and the PSG data and LM data were aligned based on the start and stop time for data analysis. After the participants woke up, the experts removed the devices from them and conducted a brief interview to confirm that there were no adverse events. Participants received compensation commensurate with their burden on participating in the study within 7 days after the experiment.

All procedures in this study were approved by the IRB, Shanghai Mental Health Center (2022-12RI).

### Devices

#### PSG

The PSG assessment for this experiment used the Natus Embla NDx (Natus Medical Incorporated, Pleasanton, California, USA) system. The EEG channels recorded in the study included F4/M1, C4/M1, O2/M1, FP2/M1, and FP1/M1, with a sampling rate of 512Hz and band-pass filtering of 0.5-35 Hz. Bilateral EOG and chin EMG were also recorded. Since the participants in the study did not have any respiratory sleep disorders, the breathing and heart rate have not been recorded in order to reduce the burden on the participants. When connecting the electrodes, conductive gel was used as a medium to connect the cup electrodes and the participant’s scalp at the F4, C4, and O2 positions. Automatic adhesive electrodes were used at other locations, including M1, M2, FP1, FP2, E2, E1, Chin1, Chin2, Chinz, and the reference point.

#### LM Sleep Recorder

The LM Sleep Recorder (EEGION Co., Ltd) used in this study was the X7-mini version, which is the lightest version in the LM Sleep Recorder series. It consists of a base box for charging and data processing, and a head patch for recording and data transferring. These two components communicate via Bluetooth protocol, and the packet loss rate is less than 0.1% when the base box is placed within 2 meters from the patch. The base box has internet connectivity and is available in both WIFI and 4G-LTE versions. The recorded data will be uploaded to the cloud server in time when the device is connected to the internet. The analysis results, which include sleep staging and other sleep variables, will be sent to the user after the online process finishes.

The head patch records frontal single-channel EEG at the FP1-FP2 and three-dimensional head movement data, which are necessary for accurate sleep staging. The head patch is connected to the forehead skin with a specially designed electrode, which contains a flexible circuit board for signal transmission and skin-friendly hydrogel in contact with the skin. Additionally, the electrode uses a hollow breathable design, which greatly reduces the burden on users and makes it more comfortable to wear.

### Data Analysis

In this experiment, we analyzed PSG and LM data from two aspects: consistency of the raw signals and consistency of sleep analysis results. Further, sleep analysis results included sleep staging results and multiple sleep variables.

Due to LM records only one channel EEG at the FP1-FP2 position, the EEG of the same PSG channel was used when evaluating the consistency of raw signals. In contrast, when evaluating the consistency of sleep analysis, all channels of PSG data were used.

#### EEG brain wave similarity

Since we cannot display the raw signal of the two kinds of data for all records, we need some indicators to objectively evaluate the consistency of the data. Follow Sterr’s work[17], we extracted the sleeping parts of each record and calculated the RSP of each frequency band, including delta (0.5-4 Hz), theta (4-8 Hz), alpha (8-12 Hz), sigma (12-16 Hz), and beta (16- 30 Hz), with the total frequency range from 0.5Hz to 30Hz. Due to differences in the recording locations and the hardware, we do not expect the RSPs of each record of LM and PSG to be strictly equal, but they should follow the same trend as the participant’s overnight sleep state changes. Therefore, we measured the consistency of EEG from both devices by calculating the pearson correlation coefficient (PCC) of the RSP in each frequency band.

To be specific, we used the fast Fourier transform to calculate the power spectral density (PSD) in each 15-second segment of all the data, and then used Simpson’s rule to calculate the RSP from PSD. Before calculating the PCC, the RSP sequences had to be aligned in time.

#### Assessing sleep stages classification performance

After collecting PSG and LM data in each experiment, the two records were processed separately to obtain sleep staging results. The LM record was uploaded to cloud server through the network, where the LM automatic sleep staging algorithm was deployed to make predictions and return the results. The PSG record was given to three sleep experts at the cooperating hospitals to do sleep staging, with N1 and N2 combined as light sleep and N3 as deep sleep, corresponding to the results of the LM sleep staging algorithm.

### PSG Sleep Staging Principles

The PSG sleep staging of adults is mainly based on the data from the F4/M1, C4/M1, and O2/M1 channels (alternatively, F3/M2, C3/M2, and O1/M2 channels can be used). Epochs are classified every 30 seconds based on the biomarkers that show in the epoch. For example, the stage of wakefulness is characterized by more eye and muscle movements, N2 sleep is characterized by spindle and K-complex waves, and N3 sleep is characterized by slow-wave oscillations[5]. Additionally, since the frequency components of the EEG differ across different sleep stages, the power spectrum of the EEG can be used to help distinguish different sleep stages, shorten the processing time.

Sleep staging and analysis are relatively subjective processes. Therefore, we extract a consensus as a ground truth from multiple sleep experts’ sleep staging results. Consensus results refer to the consistent outcome among all the sleep specialists’ agreement. When there were discrepancies among the results of multiple sleep experts for the same epoch, they would first review the epochs and discuss, try to reach an agreement about the results. If they still could not decide after discussion, the epoch would be excluded from the results. The final results would serve as the ground truth for the record.

### LM Feature Extraction and Sleep Staging

Automatic sleep staging algorithms in published works usually use 30-second epoch for classification, which is consistent with the practice recommended by AASM. However, in some scenarios, this time is too long. Moreover, numerous studies have revealed that using a different epoch length, such as 10 seconds per epoch, for sleep stage classification can also yield satisfactory results[18]. Therefore, LM’s automatic staging algorithm uses 15-second epochs primarily to speed up processing of a single epoch.

It should be noted that manual sleep staging based on PSG data uses an epoch length of 30 seconds, while LM’s automatic sleep staging algorithm uses an epoch length of 15 seconds. Therefore, for valid comparison of sleep staging results, each epoch in the PSG staging results needs to be duplicated to make sure the length of the staging results is consistent in both methods.

### LM Sleep Analysis Algorithm

#### Technology Selection

The data used for training includes 121 records of full-night sleep data collected from 60 users. Considering that this dataset is relatively smaller compared to other open-source datasets while deep neural networks typically require large amounts of data for optimal performance, and the fact that features of sleep EEG is intuitive, we opted to develop the sleep staging algorithm base on LightGBM framework[16] which applies decision tree[14] as base unit. For the input raw signals, the algorithm performs feature extraction every 15 seconds, followed by classification of sleep stages with the features input to the well-trained model. The prediction performance of the algorithm is comparable to various neural network models[13] in our dataset.

The decision tree is generated automately. After setting the parameters such as the width and height of each tree and the maximum number of iterations, the LightGBM framework automatically generates each tree and splits tree node according to its own rules. The final model is presented as a random forest. When performing prediction, each tree produces its own classification result based on the input features, and the final result is generated by the voting of all the trees.

#### Model Developing

The extracted features mainly include the following: frequency domain features, time domain features, body movement features, and meta-features[19]. Frequency domain features mainly comprise the power of various common brain waves and the ratio of power in multiple frequency bands. Time domain features include statistical features of the EEG signal (such as skewness, kurtosis, interquartile range, and zero-crossing rate), fractal dimension, as well as the number of specific biomarkers, including slow waves and spindles. Body movement features mainly involve the average body movement values in a 15-second window along the three axes and the averaged value. The amplitude of body movement is reflected by the differential values along the time dimension of the force magnitude in each direction. The larger the value of body movement along a certain axis, the faster the movement speed along that axis. Meta-features include the user’s age, gender, and height-to-weight ratio, which can help the algorithm better adapt to different populations. A total of 69 basic features were extracted from the raw EEG and body movement signals.

Furthermore, considering the periodic nature of sleep, the automatic staging algorithm also needs to take context into account in order to obtain results that are more consistent with biological rhythms. Therefore, in addition, each of the 54 basic features, except for the meta-features, is averaged over an epoch-based window. The averaging process contains two types: 1. 15-epoch window average with the current epoch as the last epoch; 2. 25-epoch window average with the current epoch as the middle epoch. After averaging, each basic feature except for the meta-features generates an additional two features. The total count of features used for training and prediction is 201. The details are shown in **Table 1**.

**Table 1.** The features used in LM sleep staging algorithm for model training and prediction.

| Feature Type | Count |  | Example |
| --- | --- | --- | --- |
|  | Before Averaged | After Averaged |  |
| Frequency Domain | 41 | 123 | Delta/Alpha Power |
| Time Domain | 13 | 39 | Skewness, Petrosian |
| Head Movement | 12 | 36 | X/Y/Z Mean Difference |
| Meta | 3 |  | Age, Sex |

### Scoring performance metrics

In order to evaluate the performance of the LM automatic sleep staging algorithm, we calculated the Accuracy, F1-Score[20], and Kappa-score of the sleep staging results against to the ground truth as evaluation metrics. These metrics are computed as follows:

1. Accuracy:

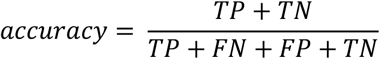

2. F1-Score:

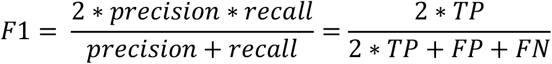

3. Kappa-Score:

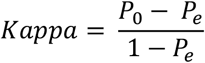

The Accuracy and F1-Score are based on confusion table, which composed of 4 classes: true positive (TP), false positive (FP), false negative (FN), true negative (TN). Giving a ground truth, accuracy provides an intuitive description of the algorithm’s probability of correctly classifying each category, while the F1-Score balances precision and recall from another perspective. The higher the F1-Score, the lower the probability of misclassification.

The Kappa-Score just evaluate the consistency between two results without a ground truth. The closer the Kappa-Score is to 1, the higher the consistency, and the closer it is to 0, the closer it is to no consistency, meaning a random relationship. Therefore, by calculating the Kappa-Score between the ground truth and the algorithm prediction, we can evaluate the consistency between them.

## RESULTS

### Raw EEG Similarity

Firstly, the direct comparison of raw signals is shown in **Figure 1**, the signals are presented separately by different stages. The four stages, each 40 seconds long, were aligned and extracted from both PSG and LM data. It’s easy to find that PSG and LM maintained consistent trends and high similarity, with biomarkers like spindles and slow waves found in the light sleep (N1/N2) and deep sleep (N3) stages, respectively.

**Figure 1.**
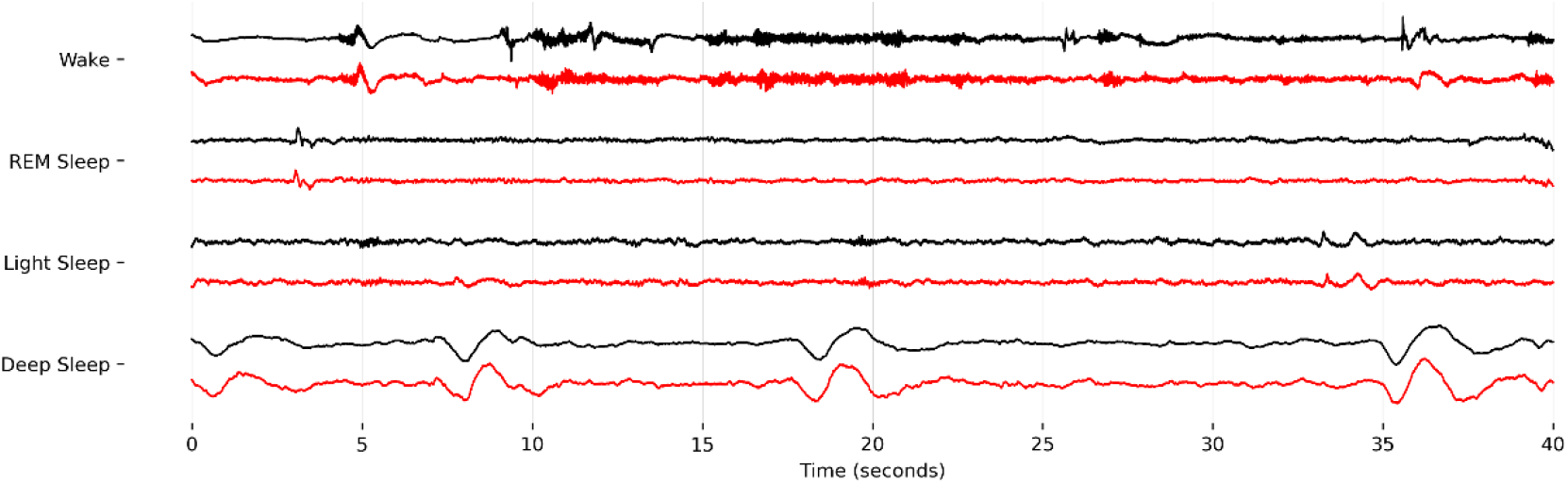
An example of the comparison of EEG signals recorded at the same time by LM and PSG during four different sleep stages. The black line refers to PSG data, the red line refers to LM data.

Secondly, the relative spectral power (RSP) of multiple frequency bands between LM and PSG can be quantitatively evaluated by the pearson correlation coefficient (PCC), as shown in **Table 2** and **Figure 2**. The following frequency bands, which are important in sleep EEG, were selected to compare: delta (0.5-4Hz), theta (4-8Hz), alpha (8-12Hz), and sigma (12-16Hz). In this part, we abandoned epochs during wakefulness in order to avoid signal changes dramatically due to body movement.

**Table 2.** The average Pearson correlation coefficient of all records collected in this study, shown in different wave bands.

| Band Name | Band Range (Hz) | Avg Corr |
| --- | --- | --- |
| Delta | 0.5~4 | 0.8816 *** |
| Theta | 4~8 | 0.7613 *** |
| Alpha | 8~12 | 0.7390 *** |
| Sigma | 12~16 | 0.8353 *** |

\*\*\*p<0.001

**Figure 2.**
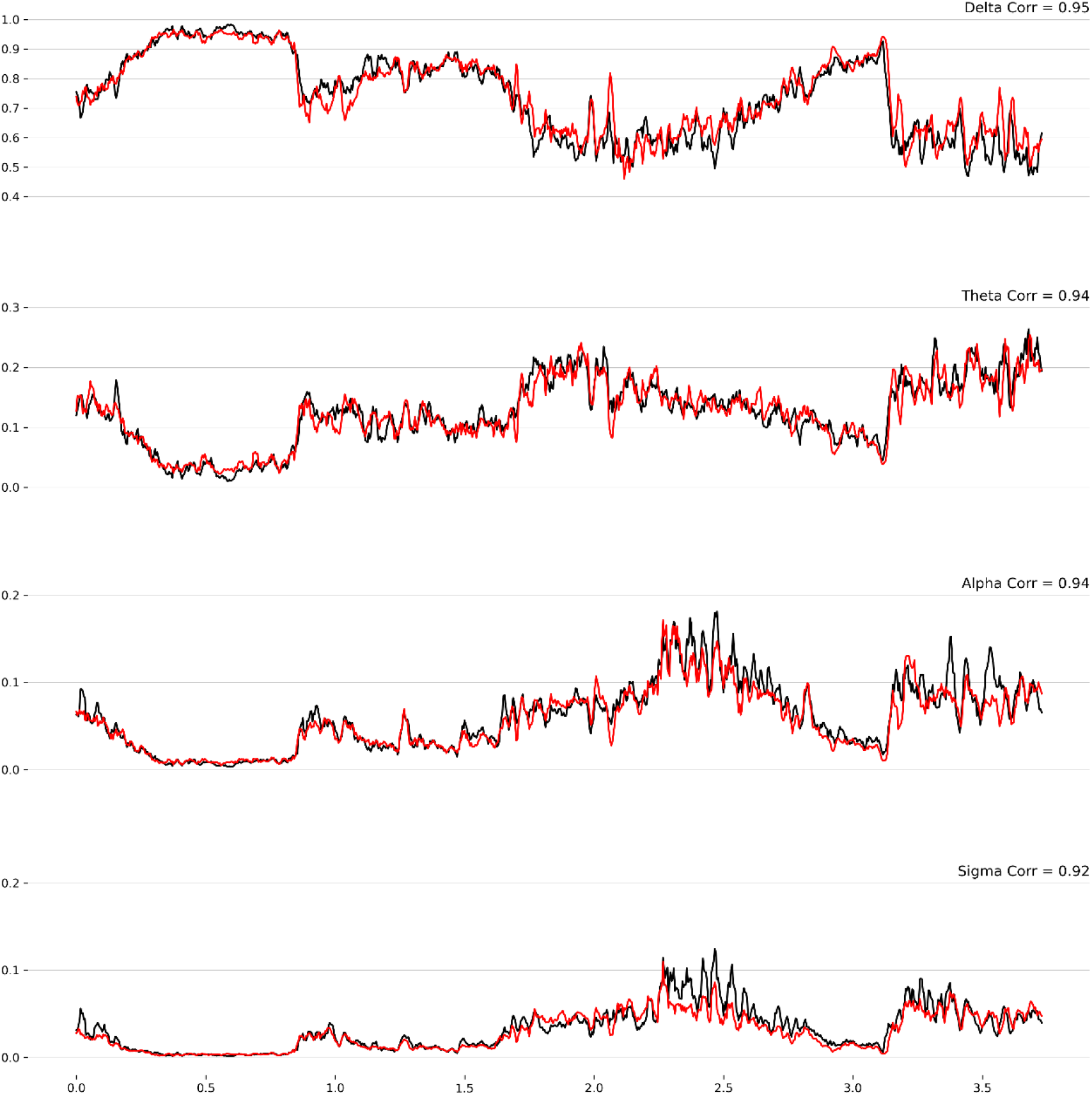
An example of the comparison of Pearson Correlation Coefficient, recorded both from sleep start and sleep end, includes multiple sleep stages. The black line refers to PSG record, while the red line refers to LM record.

The average PCC of all data for each frequency band is recorded in **Table 2**, and the PCC for all frequency bands are above 0.7, with correlations for the delta and sigma bands being above 0.8. As shown in **Figure 2**, the PCC in the delta band is often the highest because delta waves are the most prominent component brain waves during sleep, with high power and therefore high signal-to-noise ratio. This suggests that the two records of EEG have high consistency in the raw signal in terms of frequency component. Considering that the skin locations for data collection are not exactly the same and that the internal structures of LM and PSG devices are not consistent, this result is acceptable and can be considered effective.

Due to the difference in sampling rate between the two devices, the data used to plot **Figure 1** was resampled, meaning the 512 Hz PSG data was resampled to 500 Hz. This ensures that they have the same number of sampling points in the same time duration. However, resampling is not necessary when calculating the RSP, as the power spectrum can be calculated for each data according to their own sampling rates.

### Sleep stage comparision

**Table 3** shows the overall consistency between the automatic sleep staging results obtained from the LM algorithm and the ground truth. The overall F1-Score for all records is 84.03%, and the accuracy is 84.12%. The classification F1-Scores for each class obtained by the LM automatic sleep staging algorithm, from highest to lowest, are Wake (93.67%±9.49%), REM Sleep (87.23%±19.74%), Deep Sleep (82.82%±18.78%), and Light Sleep (72.10%±13.06%).

**Table 3.** Multiple sleep staging performance metrics for the LM automatic algorithm against PSG sleep experts’ consensus, each value is presented in mean ± SD format.

|  | F1-Score | Accuracy | Cohen Kappa |
| --- | --- | --- | --- |
| All | 84.03% ± 8.33% | 84.12% ± 10.31% | 78.81% ± 4.87% |
| Wake | 93.67% ± 11.14% | 96.75% ± 11.90% | 91.49% ± 12.95% |
| REM Sleep | 87.23% ± 9.49% | 93.92% ± 7.81% | 83.25% ± 17.47% |
| Light Sleep | 72.10% ± 7.61% | 86.79% ± 5.32% | 63.46% ± 9.09% |
| Deep Sleep | 82.82% ± 7.39% | 90.78% ± 11.33% | 76.53% ± 7.43% |

It can be seen in **Figure 3**, which shows the sleep stage comparison results of 4 recordings. The primary differences usually occur during the later stages of sleep, NREM sleep becomes generally shallower, and misclassification between deep and light sleep is more likely to occur.

**Figure 3.**
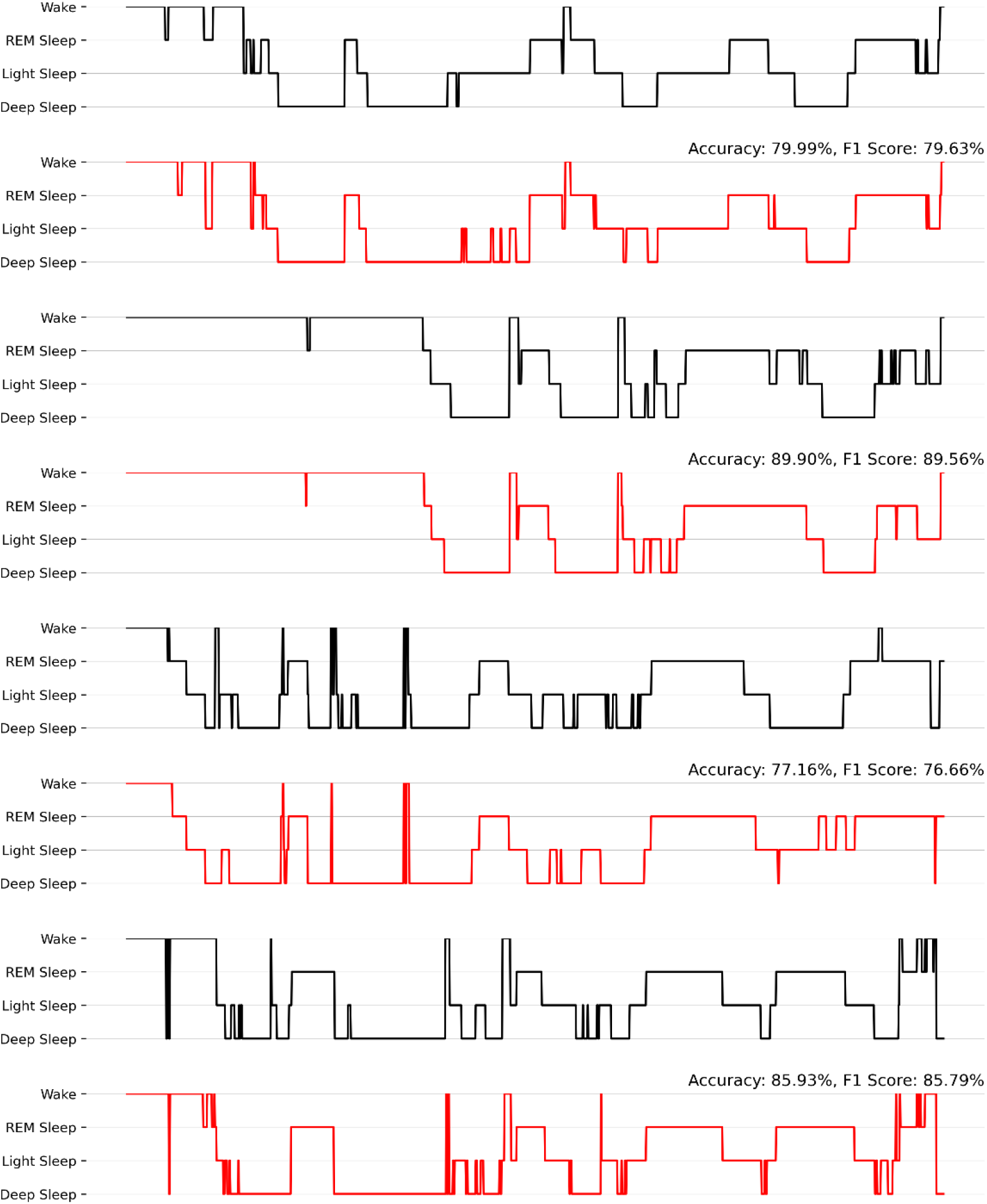
Examples of the comparison of sleep staging results. The black line refers to PSG consensus result (ground truth), while the red line refers to LM result. The evaluation metrics of each example are presented in the middle of each subplot.

The confusion matrix (**Figure 4**) provides overall statistics for each classification result. According to the matrix, light sleep is most commonly misclassified as deep sleep (18.76% probability), followed by REM sleep (10.6% probability). Deep sleep is also most easily misclassified as light sleep, with a probability of 13.38%. On the other hand, REM sleep and wakefulness have relatively good performance, with correct classification probabilities both above 90%.

**Figure 4.**
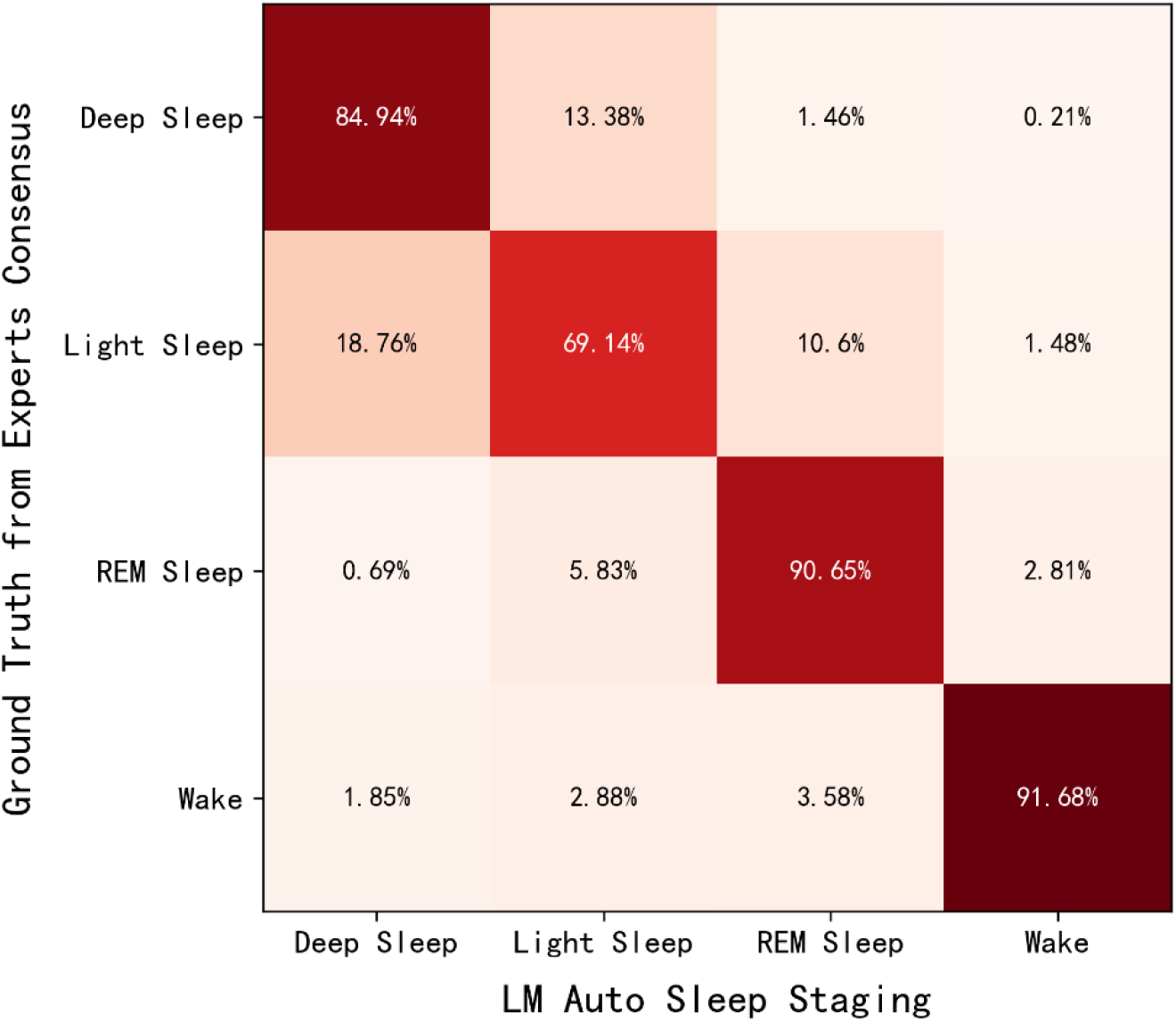
Confusion matrix for the LM sleep staging results versus PSG scoring consensuses. The value in the *i-th* row and *j-th* column represents the proportion of the *i-th* class in the ground truth being classified as the *j-th* class.

### Sleep variables comparision

We also compared the differences between the sleep variables calculated by LM automatic algorithm and those calculated by human experts for each record. These sleep variables include: 1. Total Sleep Time (TST, in minutes); 2. Sleep Efficiency (SE, %); 3. Sleep Onset Latency (SOL, in minutes); 4. Wake After Sleep Onset (WASO, in minutes). Since we have already showed the comparison results of sleep staging in detail earlier, we do not discuss the variables that can be directly calculated from the sleep staging results, such as the proportion of each sleep stage.

Due to the small sample size, Bland-Altman plots were used to show the difference of each sleep variable, each sample point refers to a whole night sleep record, shown in **Figure 5**. The range of mean±1.96 standard deviation was used as the confidence interval. Since there is a ground truth (PSG manual result) among the two results, we chose the PSG manual result as the abscissa for each sample point, rather than the average of the two results. It can be seen from the plots that none of the sample points exceeded the confidence interval for the 4 variables, indicating that there were no significant differences between the sleep variables calculated by LM and those calculated by sleep experts.

**Figure 5.**
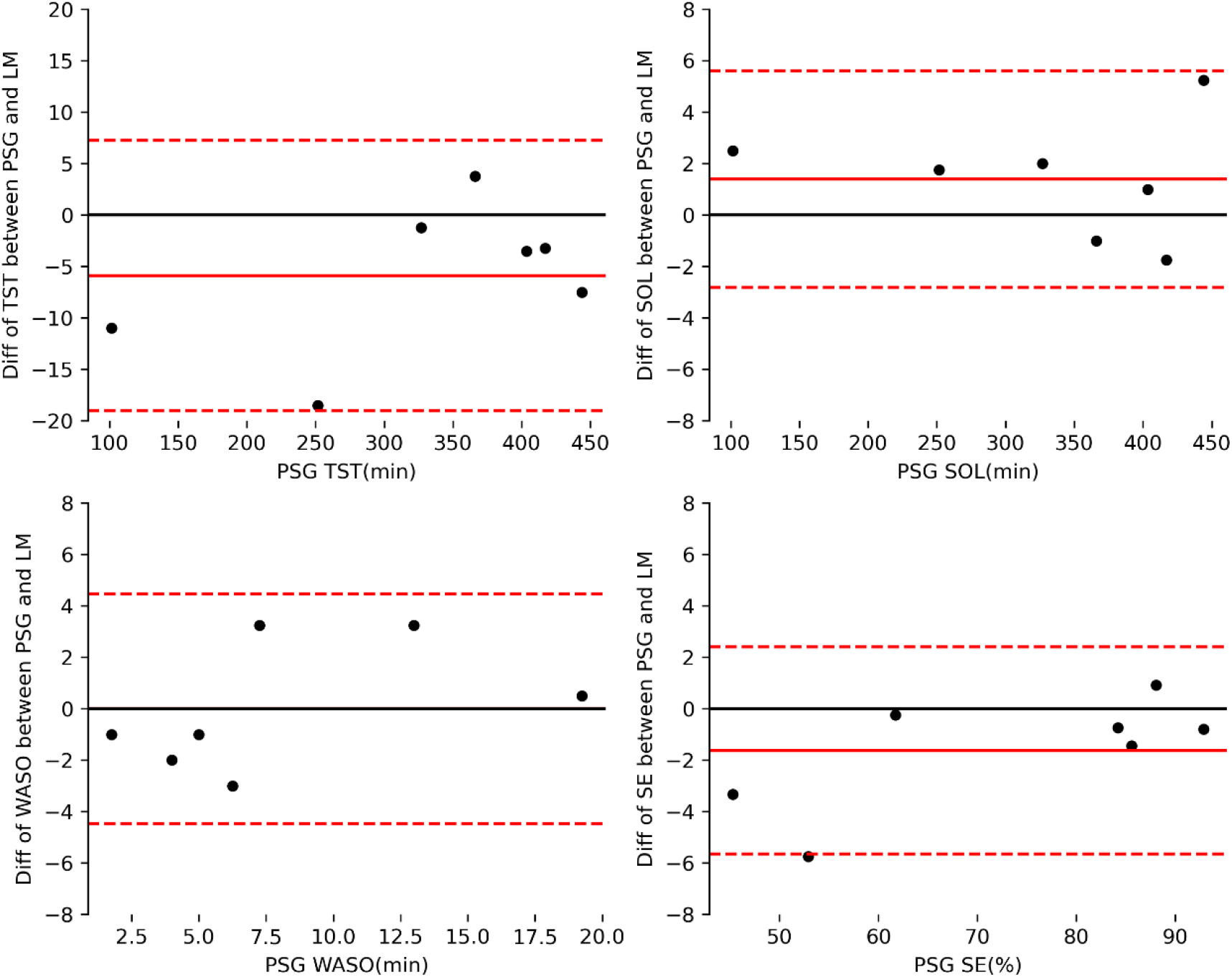
Bland-Altman plots for 4 sleep variables generated separately by LM and human sleep experts.

## DISCUSSION

In this study, we mainly verified whether LM wearable EEG devices could serve as a partial replacement for PSG devices in the home environment, specifically in terms of EEG signal collection and sleep analysis. The direct comparisons of EEG signals from the two devices indicated that LM provides similar results to PSG. We also found that the accuracy of sleep staging produced by LM is comparable to sleep scoring using PSG data. Our results indicated the possibility of applying LM as a sleep measurement tool at home.

EEG signals of LM records generally consisted with PSG records and sleep staging results between LM automatic algorithm are comparable to sleep experts based on PSG. The performance on stage Wake is the highest, the reason for this may attribute to the EEG during wakefulness has the most complex brain wave components, with high power both in high and low frequency components, which make it easy to tell apart from other stages. The accuracy of light sleep is relatively poor, which has been observed in previous studies [8][21]. This may be due to the characteristics of light sleep includes both N1 and N2 stages, which are relatively less obvious, as their spectral features are similar to those of N3 sleep. Therefore, light sleep is often misclassified as deep sleep, or vice versa. Research has indicated that in a feature space composed of multiple sleep features, sample points of the N1 stage often mix with other stages and are difficult to form independent clusters, which suggests that the features of the N1 stage are not distinct[21]. Compared to the poor performance of light sleep, the detection of deep sleep is relatively high. One possible explanation is that N3 sleep is the deepest stage of sleep, with the highest proportion of low frequency components and the lowest proportion of high frequency components, makes it easy to distinguish. The higher accuracy of the deep sleep allows us to try some more intriguing endeavors, such as enhancing slow waves through phase-locked pink noise. Although this phenomenon is easily reproducible in laboratory settings[22], it remains challenging to work stably on wearable devices. Nonetheless, we have made significant progress in this regard and plan to publish the work in the future.

Automatic staging algorithm plays an important role in sleep staging classification of portable EEG devices. Sleep experts who refer to the AASM standard mainly focus on EEG biomarkers, in the current study, LM’s automatic staging algorithm mainly uses power spectral features, which are easier to distinguish the transition of sleep stages throughout the night, especially the transition between REM and NREM. In addition, the counts of slow-wave, spindle are also calculated as features to distinguish between deep and light sleep, which is consistent with AASM’s principles. Therefore, in comparison to other deep neural network-based methods, the LightGBM framework used in this study exhibits superior interpretability.

In terms of sleep variables, LM automatic sleep analysis algorithm achieved similar results to those obtained by human experts based on PSG for TST and the proportion of each sleep stage, thanks to the higher accuracy of sleep staging. The algorithm tended to overestimate WASO, and this may be due to the lack of information on eye movement and muscle activity in the LM data, which makes it easier for the algorithm to misclassify movements such as turning over as awakenings. To address this issue, future work may shorten the epoch length used to 10 seconds or even 5 seconds. It may be possible to detect movements such as turning over more accurately, reducing their impact on the context and minimizing false detections of awakenings[23].

Compared to clinical PSG devices, LM has the advantages of being user-friendly and highly acceptable. On the other hand, compared to other devices such as smart watches, LM offers more scientific and precise sleep monitoring through the frontal EEG, making it an excellent choice for individuals who want to understand their sleep quality and for doctors who want to track their patient’s sleep at home. However, some potential limitations need to be discussed when applying LM to access sleep. First, the sample was small and only included people with no or mild symptoms of insomnia. Further studies should run on large samples, and recruit individuals who suffer from diverse sleep disorders to verity the LM device. Second, the LM sleep analysis algorithm in this study has not developed to be adaptive to specific users. Human sleep characteristics follow overall statistical patterns, there are slight differences among individuals. For example, the average spindle number of N2 period varies among individuals of the same age group and changes with age for the same individual[24]. Human sleep experts can fine-tune the analysis results based on the specific sleep characteristics of each patient, resulting in higher compatibility between the results and the patient’s actual condition. Thus, in future work, LM’s automatic algorithm should incorporate the functionality of extracting personalized features of users to generate sleep analysis results that are more compatible with specific users. Last but not least, in the study we simulate the home environments in the hospital center while situations may be more complex in real life. Thus, further studies should consider extra factors at home when using the LM device.

## CONCLUSION

In this study, we have demonstrated two main functions of the lightweight wearable device, LM, by comparing its data and analysis results with those of clinical PSG records. These functions include: 1) the ability to record high-quality frontal single-channel EEG that is comparable to PSG, and 2) the ability to automatically perform sleep staging and sleep variable calculations with an accuracy level that is comparable to that of human sleep experts. The lightweight and easy-to-use nature of LM makes collecting sleep data easier, which makes constructing a big sleep data platform possible. In the future, it is possible to conduct various specialized sleep health research and treatment based on specific populations and groups.

## Data Availability

All data produced in the present study are available upon reasonable request to the authors.

## ACKNOWLEDGMENTS

We would like to thank Shanghai Center for Brain Science and Brain-Inspired Technology for providing sleep monitoring rooms.

We also would like to thank the Shanghai Quanlan Technology development team for their hard work on the LANMAO Sleep Recorder both in hardware and software.

## DISCLOSURE STATEMENTS

This study was supported by Shanghai Quanlan Technology team.

All data used in the present study are available upon reasonable request to the authors.

